# Stakeholders’ perspectives on the feasibility of adopting a Healthy Nail Salon Recognition Program in Philadelphia: A qualitative study

**DOI:** 10.1101/2022.09.15.22280008

**Authors:** Duong T Nguyen, Chau Nguyen, Jessie Pintor, Tran B Huynh

## Abstract

**Background:** The California Healthy Nail Salon Recognition Program is a statewide initiative to incentivize nail salons to adopt safer practices such as use of safer nail products without certain harmful chemicals, installation of ventilation systems, proper personal protective equipment use, and staff training. This public policy intervention is in response to the call to protect nail care workers, mostly women of color, who bear disproportionate burden of chemical exposure at work. Because there is interest from the community to adopt similar program in the Greater Philadelphia region, we conducted this formative research to document stakeholders’ perspectives on the feasibility of adopting the Healthy Nail Salon Recognition Program in Philadelphia.

**Methods:** We conducted semi-structured interviews with a purposive sample of 31 stakeholders in Philadelphia in 2021. Using the Consolidated Framework for Implementation Science as our theoretical framework, we developed the interview guide and analyzed the data using qualitative research method to identify key facilitators and barriers.

**Results:** Key facilitating themes were perceived need and benefits of program to improve workers’ health and working conditions, and willingness of stakeholders to leverage their organizational resources. Barriers included perceived high cost and time commitment from salon owners and employees, lack of funding and implementation leaders at the city government, community members’ willingness to be visible and advocate for the program affected by the stigmas of being immigrant workers, fear of interacting with authorities, as well as the impact of COVID-19 pandemic. Our results **suggest** successful adoption of the Healthy Nail Salon Recognition Program in Philadelphia will require outreach within the community to raise awareness of the benefits of the program and close partnership with community-based organizations to facilitate mutual understanding between the authority and the ethnically diverse nail salon communities.

## INTRODUCTION

Low wage, precariously employed, minority nail care workers is an underserved community in the United States (US) (Sharma et al.,hh 2018). Despite the documented harmful respiratory, neurological, and reproductive health effects from chronic exposure to low levels of chemicals from nail products and other work-related hazards in this worker population across the country (Dang et al., 2021; Huynh et al., 2019; LoSasso et al., 2002; Ma et al., 2019; Nguyen et al., 2022; Quach et al., 2008; Quiros-Alcala et al., 2019; Roelofs et al., 2008; Siegel et al., 2021). Limited oversight at the federal level combined with patchwork of laws at the state level to regulate toxic chemicals in personal care products and cosmetics have resulted a plethora of harmful products available to the consumers (U.S. Food and Drug Administration, 2022, Campaign for Safe Cosmetics, n.d.). Alternative safer products tend to be more expensive and limited in availabilities. With the lack of comprehensive federal oversight of big chemical manufacturers and distributors of nail products, local and state governments turn to regulate the workplaces, putting the responsibilities of reducing chemical exposures on small business owners, the workers, and the consumers. Women of color with disadvantaged backgrounds bear the brunt of the chemical exposure burden. Currently very few local and state governments have been able to develop public policies to address this occupational health injustice (*Bill Text - AB-2125 Healthy Nail Salon Recognition Program*., n.d.; *Healthy Nail Salons - Local Hazardous Waste Management Program in King County*, n.d.; New York State, n.d.; Seller et al., 2019)

In California since 2010, with the help of the California Healthy Nail Salon Collaborative, several local health departments have implemented healthy nail salon recognition programs (HNSRP) to incentivize salons to adopt best practices.(California Nail Salon Collaborative, n.d.) Nail salons which meet the program criteria are certified as “healthy nail salons”. These criteria include use of safer nail products without certain harmful chemicals, installation of ventilation systems, proper personal protective equipment use, and staff training.(*Healthy Nail Salon Recognition Program*, n.d.) The success of these local programs has led to the passage of the CA Assembly Bill AB2125 that requires the CA Department of Toxic Substances Control to publish guidelines for local cities and counties across the state of CA to voluntarily implement the program (*Bill Text - AB-2125 Healthy Nail Salon Recognition Program*., n.d.). A pilot program evaluation of the HNSRP conducted in 2012 in San Francisco (the first county to adopt the program in CA) suggested early evidence of positive impact of the program on the salon environment and the workers (Garcia et al., 2015). In this ‘natural experiment’ study, the research team conducted personal air monitoring and surveys assessing knowledge and behaviors of workers from six participatory salons (intervention group) and five salons that did not participate in the program (control group). They found decreases in levels of toluene and total volatile organic compounds, as well as increase in knowledge and self-reported positive practices in the participatory salons compared to the control group.(Garcia et al., 2015)

The Greater Philadelphia region is one of the most populous US metropolitan areas that has high concentration of Asian American communities and minority-owned nail salons. While there has been documented health needs locally (Freeland et al., 2021; Huynh et al., 2019; Ma et al., 2019), public policy to address the issue is lacking. There is interest from the community to have the city of Philadelphia to adopt similar program which could potentially serve two purposes: 1) incentivize salons to improve the work environment, and 2) an opportunity for the city and community-based organization to work together to amplify outreach efforts within the diverse communities in Philadelphia. The purpose of this study is to document stakeholders’ perspectives on the feasibility of adopting the HNSRP in Philadelphia. Findings from our study could be useful for local policy makers and public health authorities to assess their programs and functions and for community organizers to strategize their next steps to advocate for programs that meet the needs of the community.

## METHODS

### Theoretical framework

We used the Consolidated Framework for Implementation Research (CFIR) as the basis for the design of this study. The CFIR incorporates a suite of constructs from the literature that have been associated with effective program implementation. The framework includes five domains: intervention characteristics, outer setting, inner setting, characteristics of individuals, and process and constructs within each domain. The website dedicated to the dissemination of CFIR offers numerous resources for implementation science practitioners to use in their evaluation study (*The Consolidated Framework for Implementation Research – Technical Assistance for Users of the CFIR Framework*, n.d.). In the context of this study, we selected constructs related to organizational readiness to qualitative gauge stakeholders’ perspectives on the city’s level of readiness for such a program (Kononowech et al., 2021). This type of formative research, conducted prior to the development a program, could provide useful information to program planners and community organizers. We used CFIR construct guide both creation of the questionnaire and for data analysis. The main questions for the interview were as follows:

> Do you think the program is needed in Philadelphia? Can you please explain your rationale?
>
> From your perspectives, what do you think will help support the implementation of the HNSRP in Philadelphia?
>
> What do you think are the potential barriers for implementing the program?
>
> How do you see you or your organization could contribute to the implementation of the HNSRP in Philadelphia?
>
> Is there anything else you want to share with me regarding your thoughts on the HNSRP?

### Participants

We conducted semi-structured interviews with a purposive sample of 31 stakeholders in the Philadelphia area that included staff members in city and government systems (n=4), public health experts (n=2), community leaders (n=2), non-governmental organization (NGO) community outreach workers (n=2), cosmetology instructors (n=2), nail supply store owners (n= 3), nail salon owners (n=6, including 2 who are also cosmetology instructors and 1 who also owns a nail supply store), and nail salon workers (n=14, including 1 who is also an outreach worker). The Vietnamese nail salon worker community was the focus for this research due to the high percentage of Vietnamese-owned nail salons in Philadelphia.

### Data collection and analysis

We identified potential interviewees from publicly available online information on governmental websites, referrals from the community advisory board and in-person outreach. We recruited participants via emails (n=7), phone calls (n=18) and in-person visits (n=6). The Vietnamese nail salon worker community was the focus for this research due to the high percentage of Vietnamese-owned nail salons in Philadelphia. We conducted ten interviews in English and 21 in Vietnamese.

After agreeing to participate, participants received either an electronic or a physical package which included basic information of the California HNSRP, how to participate and the criteria to be certified as a healthy nail salon. They also received the interview questionnaire in advance. All phone (n=25) and in-person interviews (n=6) were recorded, translated into English if needed, transcribed, de-identified and stored in a secure digital folder until the analysis.

The de-identified transcripts were then coded using NVivo 12 by two independent coders. We identified themes using the CFIR to identify patterns of facilitators and barriers. After the data analysis, we generated a bilingual preliminary report and sent to all participants for feedback and validity check for the data interpretation.

The datasets and codes generated and analyzed for this study are available from the corresponding author on reasonable request. This project was reviewed and approved by Institutional Review Board (IRB) of Drexel University

## RESULTS

Figure 1 summarizes key facilitators (green), barriers (red), and themes that can be both a barrier and facilitator (yellow) to adopting a HNSRP in Philadelphia in the context of the CIFR framework.

**Figure 1:**
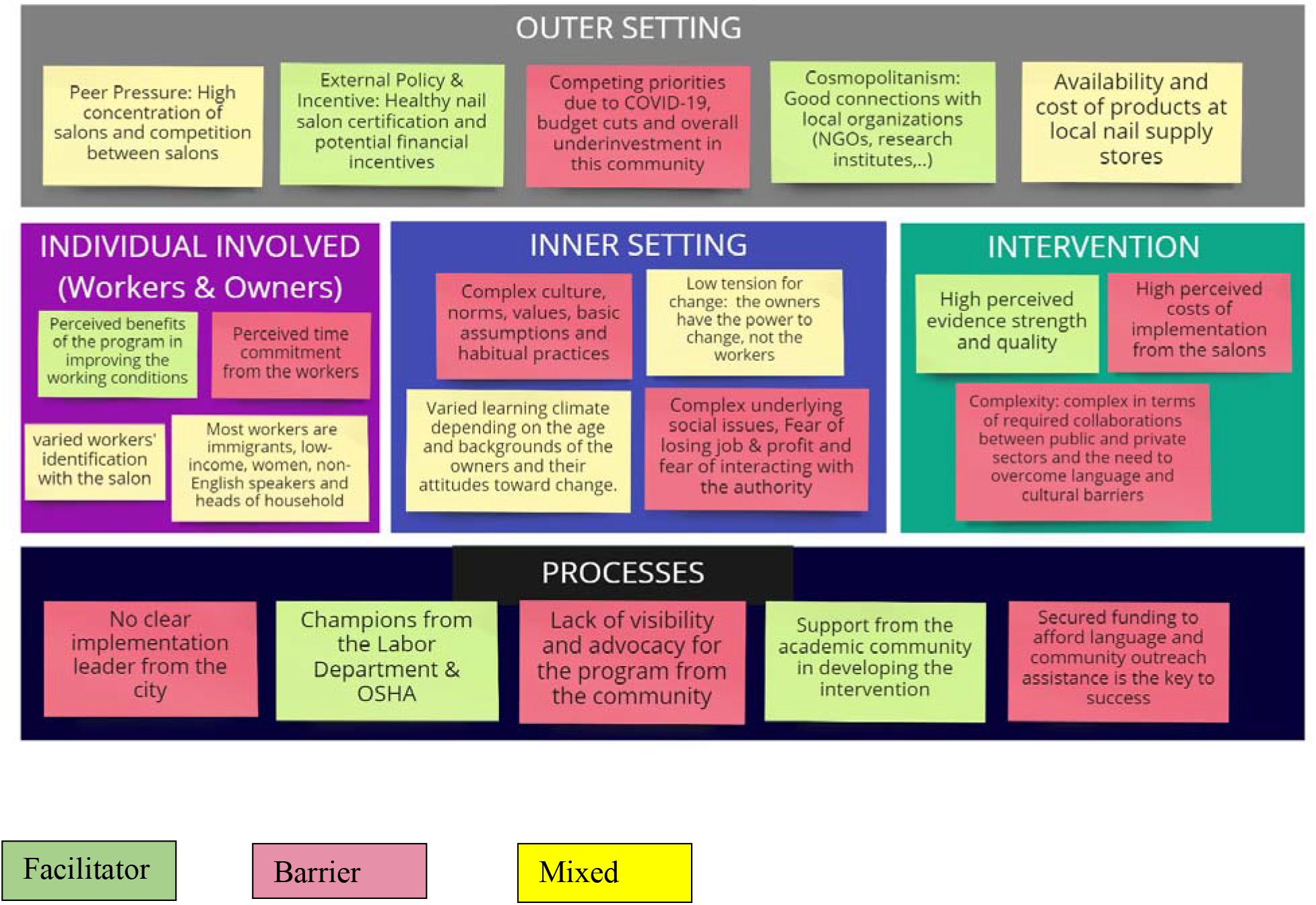
Summary of themes in the context of the CIFR framework. Potential key facilitators are in green, barriers in red, and themes that can be both a barrier and facilitator in yellow.

### Key facilitators

#### 1. Perceived benefits of the program to improve health and working conditions

All participants believed the program would raise the awareness of work-related health issues in their community and ensure workers are professionally trained and protected. Many interviewees expressed concerns about nail salon workers and owners currently not having access to accurate safety information and training including worker’ rights due to language barrier. From the community’s perspective, this program would fill the gap in the current cosmetology training and development and ensure the workers’ rights are protected A director of a local community organization serving many Vietnamese nail salon workers and owners said, “I know that nail salon workers who work with many chemicals may not know their rights in the workplace. Many times, it’s because they’ve never come across the information, or they may have heard some misinformation.” Many salons worker and owner participants also emphasized the importance of on-going safety training as the nail salon industry is the major source of income for many Vietnamese families and most workers are chronically exposed to chemicals for long hours every day. An owner said, “[The program] helps protect the health of nail workers. They work long-term, connected for a long time at work. They work every day, every week, from year to year, so health is especially important.”

From the state and city’s perspectives, the program would support them to do their job by ensuring the correct information can reach the community and workers know their rights and the process to make a complaint if needed. It has been especially challenging for them to connect with the community due to language barrier. One OSHA (Occupational Safety and Health Administration) official said:

> The workforce has limited English proficiency and limited information about what we can do for protection. They might not know what the process is and what their rights are to make that complaint, but as you know in Philadelphia there are a lot of salons… There’s a good amount of work and workers that would need that intervention to make sure that they have proper safety measures at the workplace.

Moreover, most participants believed the program would incentivize employers to improve the work environment and being a recognized “healthy nail salon” can also have a positive impact on the salon’s public image and attract more customers. One owner expressed: “It helps the owners protect their workers better and protect the customers and the environment.*”*.

#### 2. Willingness of some stakeholders to leverage their organizational resources to support the adoption of the program

Most participants (OSHA inspector, city staff members from the Department of Labor, community leaders and NGO directors as well as nail salon owners and workers) endorsed the program and expressed interest in being involved in the adaptation and implementation of the program in Philadelphia. All the interviewees believed the program required hand in hand collaboration between the city and the community. When asked about how each stakeholder can play a role in making this initiative happen, many respondents believed either the city’s health department or labor department could take the lead. One city official expressed, “Philly is a big union town, so worker rights are very well understood. The labor office is where we should start.” The director of a grassroot organization for working-class Asian communities said:

> It would be great if our organization can be part of [the program]. Let’s say the Office of Labor receives city funds to hire a staff member that supports the certification of this program. They could work with community-based organizations like us, which lead to putting together a training program that nail salon owners and workers must go through and commit to implementing these steps. And then you can get city recognition as a certified healthy nail salon. Once that certification happens and the Office of Labor needs to periodically check in to ensure that the practices are in place… and that could also happen with the support of a community-based organization that has established those relationships with those new clients.

#### 2. Perceived as a necessary step to improve the underserved salon community

Most interviewees acknowledged the need for investment into the nail salon immigrant worker community and perceived the program to be a necessary step to create trust and partnership between the city and the community. From the public health’s perspective, the program was believed to have a powerful impact in facilitating better understanding about the issues the community is facing and thus help address the current health disparities and social inequity. A city’s public health expert said:

> [The program] will help us better understand the health disparities and gaps and negative health outcomes and negative working conditions. What are the biggest issues that are impacting this community and how are they impacting them? It will help us improve health outcomes and hopefully address the health disparities in the nail salon worker community in Philadelphia. I do not think anyone else is going to do it.

This initiative can also the way for future public health programs and workers’ rights initiatives, as well as community development and empowerment projects. A city official said, *“*The program can assist with community engagement and help the nail salon community feel like they are an important part of the larger Philadelphia community and help people feel like they are prioritized.*”* A state official also said, “It’s important for the city of Philadelphia, and for the state to know how diverse our communities are and for us to develop programs that can impact that through workplaces, where people get their livelihoods, and we want these communities to be safe at our workplaces.” One public health expert said, “The program might help members of that community to become more empowered to take charge of their own health or to be more or to be advocates, or to be more engaged in community level work, community policy, things like that.” This also translates into the development of projects to empower immigrant-owned, woman-owned small businesses and uplift the entire working-class, immigrant community.

### Key barriers

#### 1. Perceived high cost and time commitment from the owners and workers

Costs and time commitment are the biggest barriers for many owners and workers to participate in the program. Most nail salon workers interviewed work 5-6 days a week, 9-12 hours a day and reported not having time for continued training and professional development. They said they would be much more likely to participate in the training if it were paid. One salon worker expressed:

> I think first is income. If people have enough money to provide for their family, they will have the peace of mind to work. But now they can’t even make the basic income, how can they think about anything else? If they make enough money, they may want to talk with you. But now their income is too low, all they care about is compete for customers, they would not have time for you.

Several salon owners acknowledged the benefits of the program and willingness to change their salon’s environment and working habits but considered the costs to upgrade their nail salons to meet the certification standards to be a prohibitive factor. They expressed concerns that switching to safer products with less harmful chemicals and upgrading the ventilation systems would cost them a lot of money. Most owners said that their family business had already been operating at a very narrow profit margin. Therefore, increased costs could be detrimental to their businesses. An owner expressed: *“*It will be hard for us because we will have to pay more and take care of more things and it will be more restrictive for us to do business.*”*

#### 2. Funding to afford language assistance and community outreach

The program was perceived by some stakeholders as “complex” given language and cultural differences that need to be overcome. Some said it could come across as “too niche to be funded” given the issues in this industry not being well-known at the city level. A director of a community organization said, “I think the service industry in general is overlooked by the city… There’s very, very little attention paid to language access. If there was ever a recognition program that was rolled out with language access, primary language access must be at the forefront.” A state official also expressed concerns around funding sources for this initiative:

> A potential barrier is that it would seem so specific that it will not be impacting a larger workforce that people will feel like it is too niche to fund. It is about placing the program and how it permeates various aspects of the Philadelphia economy. It would be helpful for people to understand the importance of the program situated within the bigger context.”

Others disagreed that the program is “niche to be funded.” One participant said:

> It’s not that the program is niche to fund, but I think there got to be larger education around the nail salon industry, who owns it, how it operates and the work safety standards that need to be implemented or discussed. It’s an issue with education around this workforce at the Philadelphia level in terms of city council and the city offices to learn about the importance of this. It would be good to analyze how these programs came to be in terms of funding, both at the city level but also at the state level. It will be good to present what happened in California with Santa Monica and San Francisco to try to figure out how other states support the implementation of this and having that history will help inform some of the State senators and the House representatives.

Language assistance and partnerships between the city and community organizations need to be established at the forefront for the program to be successfully implemented. On-going financial and technical support is also needed at every step of the process of planning, implementation, and revaluation.

#### 3. Buy-in and visibility from the nail salon community

Policy expert participants emphasized the importance of community’s visibility in pushing forward the adoption of the program at the city level. One city official expressed:

> I think once you’ve gotten those numbers together, build some momentum behind those numbers, increase the visibility of the program, hold town halls or community hearings or go to City Hall. Visibility is going to be important, but it can be difficult for people to step out in the spotlight. Data along with people showing up to be visible will be helpful in getting people to pay attention.

However, the community’s buy-in for the program was affected by an overall lack of awareness of the program. A community organizer said, “The barrier for this program is it’s too new so the owners may not be willing to be open-minded. They may understand this issue is important, but the owners and workers are already used to their own ways of doing things. They do not want to change. They don’t think it’s important enough for them to participate.”

The community’s willingness to be visible and advocate for the program was also affected by the stigmas of being immigrant workers, fear of interacting with authorities and lack of trust in the government, as well as the impact of COVID-19 and anti-Asian sentiment. A community leader said, “You’re talking about an industry that is mostly comprised of immigrant workers and immigrant small business owners. There are a lot of stigmas to show up to those kinds of things and a lot of mistrust.” Another community worker mentioned the fear of interacting with the authority eloquently through what he witnessed with his mother, a nail salon worker. He said:

> You know, my mom is a nail salon worker. All her friends are. When we moved here from Vietnam, we know that is the only thing. If you are a woman and you are Vietnamese immigrant, that is one of the three things you can do. The other two are working in a restaurant or working in a factory, where any of those spaces are also equally exploited. My folks do not speak English and immigrants are afraid of stirring waves and contacting with authority. They just put up with it. My mom was exploited her whole life. But every time I am like ‘you should put a complaint’ and she is like ‘no.’ She does not want to make waves. She does not want to be the one who snitched out… It is just a big shame to have it happen to her in the first place.

All the interviewees from the community emphasized the importance of making the intention of the program clear from the beginning and educating the community on the benefits of the program to improve buy-in and encourage participation. One nail salon worker said: “You should make it clear that this program is designed to help them and the community, not to report anybody.”

#### 4. Lack of implementation leaders/champions within the city departments

Despite the endorsement of the program from many stakeholders there was no clear consensus on who should be the implementation leader of the program. This is a critical missing piece because the program’s transdisciplinary nature requires an implementation leader to act as a liaison to bring different city departments together to work with the community. According to the interviewees, one of the reasons for this lack of clarity is because city departments, such as the labor department, the health department, the Licensing & Inspection department, often work in silos with a narrow margin of funding on their own, which makes it challenging to create and manage new collaborative projects. In order address this issue holistically, these departments to work together and community partners can organize to force the city to pay more attention to these issues. Moreover, the interviewees emphasized the importance of enrolling the “actual people” at the city level on boards to be the champions for the program. One city official said, “It depends on who is leading. Philly is a town where it is less about the organization or the name of the organization and more about the actual person. It sucks because then, if the person leaves, there is really no permanence. It can be excellent one year and then not the next year.*”* Interviewees also expressed concerns about the sustainability of the program and agreed that one of the city departments needed to house it as an official program, where they could allocate ongoing staff time and resources.

## DISCUSSION

The purpose of this study was to bring various stakeholders’ voices to the discussion about potential public health policies to improve the work environment for nail salon workers, a community traditionally underserved in Philadelphia and in the US. One of the key barriers was lack of buy-in and visible advocacy from the community, which stems from lack of awareness of the program and fear of stigmas being immigrants. Most nail salon workers are immigrants and refugees (Sharma, 2018) who are not familiar with the legal system in the US and are afraid of interacting with the authorities. They are especially cautious when it comes to programs that require monitoring and reporting for fear of causing trouble with the authority and leading to more restrictive regulations, despite being at high risk of work-related illnesses. This visibility issue has been well documented in other public health and occupational projects working with immigrants from Latino and other Asian communities (Eggerth & Flynn, 2010; Morey, 2018; Wallace & Young, 2018). More education and outreach within the nail community is needed to raise awareness of the issues and encourage the community to be visible and advocate for policies suitable for their needs. Community-engaged participatory research, especially pilot programs where nail salon workers and owners can participate and learn about the issues and the HNSRP, are beneficial to build trust and raise awareness of the program in the community.

Another barrier was lack of government funding. This partly stems from lack of awareness of the issues at the city level and overall underinvestment in this community. Public health literature refers to this phenomenon as “structural invisibility,” society’s privileging and paying more attention to some social group over others, or simply “you cannot fix what you don’t see.” (Flynn et al., 2021) Structural invisibility manifests in lack of attention paid to language assistance and community outreach, leading to the absence or lack of surveillance data on this group of minority workers, which filters the “reality” that institutions see, what they consider as important and how they allocate funding (Flynn et al., 2021; Rodriguez-Lainz et al., 2018). It is important to reframe the narrative and recognize the issues that nail salon workers are facing are a part of larger inequities experienced by immigrants and BIPOC (Black, Indigenous and People of Color) communities, women, and small business owners, as well as working class in Philadelphia. More advocacy at the city level is needed to reveal this blind spot and bridge the gap between the authorities and community. It’s important for organizations and governmental agencies to reassess their processes and develop the institutional capacity to work with this increasingly diverse workforce more effectively.

The program was also perceived as providing better strategies for participatory nail salons to advertise for their business and attract customers. Future research on social marketing could be explored to gain insights into how HNSRP can impact the public image of nail salons and into ways to support salons to leverage this as a marketing tool. According to many salon owners and workers in the study, local nail supply stores also play a critical role in the success of the intervention given that they are the source of most chemicals and safety products. Outreach and collaboration with local nail supply stores may also facilitate the program uptake.

Perceived challenges related to cost to buy more expensive alternative products, implement engineering controls, or time commitment for staff training are not unique to nail salon industry but well-documented challenges for small businesses in general (Black et al.,1993, Honan et al., 2022; Moutray, 2009). Like many other businesses, nail salon small business owners and their employees were highly impacted by the COVID-19 pandemic with little limited support (Honan et al., 2022; Sharma, P. et al., 2022). Both salon owners and workers reported difficulty applying for pandemic relief fund such as the Paycheck Protection Program and other small business loans for owners or unemployment benefits for workers (Herrera et al., 2020; Herrera et al., 2021; Sharma et al., 2022). These reports underscore the need for public policies to support small businesses and their employees, with an emphasis of language justice for non-English speaking communities as these small businesses are the economic backbone of many new immigrant communities in the US.

Our study has several limitations. The first limitation is the inclusion of stakeholders who speak English or Vietnamese only. While Vietnamese American make up a large majority of the very racially and ethnically nail salon workforce (Sharma et al., 2018), salon owners and workers who are not Vietnamese Americans may have very different lived experiences, needs, and perspectives on this program. The second limitation is the absence of key stakeholders such as the Pennsylvania State Board of Cosmetology and License & Inspection due to their non-responding to our invitation. Another key stakeholder are the consumers. A survey of 650 consumers in LA County conducted by the California Healthy Nail Salon Collaborative found 85% would support a program that worked with salons to become a “Healthy Nail Salon” (internal report shared by Lisa Fu, California Healthy Nail Salon Collaborative). Our future work with the local community organizations will survey consumers in the Philadelphia area as this would provide critical information for advocacy purpose. Lastly, our snowball sampling through referrals may result in participation and volunteer biases and the small sample size, while suitable for in-depth qualitative inquiry, may limit generalizability of research findings.

Despite these limitations, the strength of our study is the diverse voices that we were able to bring together to shed light on ways stakeholders could work together to promote health and well-being of an underserved worker communities through public policy. Local policy makers, public health practitioners, and community organizers may find our findings useful to inform their next public health programming and/or advocacy strategies. While our study’s focus is in Philadelphia, various aspects of our findings may have relevance for other municipalities across the United States that may contemplate the adoption of similar voluntary programs to meet the needs of the nail salon workforce within their states and preserve the economic stability of small businesses. This type of study might be replicated in other geographic locations to better understand the local political landscape and the level of engagement from community-based organizations to address the issue in a more sustainable way.

## CONCLUSIONS

We used the CFIR framework to identify key facilitators and barriers to implementing the HNSRP in Philadelphia. The success of this program and many other community-engaging public health interventions, depends on the awareness of complex underlying social factors that influence the community’s willingness and capacity to participate, effective collaborations between various key players in the public and private sectors, relationship-building between the state and the communities, as well as secured funding for language assistance and community outreach.

## Data Availability

All data produced in the present study may be available upon reasonable request to the authors

## REFERENCES

Bill Text—AB-2125 Healthy Nail Salon Recognition Program. (n.d.). Retrieved May 11, 2021, from https://leginfo.legislature.ca.gov/faces/billNavClient.xhtml?bill_id=201520160AB2125

Black DA, Barron J, Berger MC. (1993) Job training approaches and costs in small and large firms. (SBA-6640-OA-91). Lexington, KY: University of Kentucky. Available at https://permanent.fdlp.gov/websites/www.sba.gov/advo/research/rs135.html. Accessed 25 August 2021

California Nail Salon Collaborative. (n.d.). Healthy Salons. https://cahealthynailsalons.org/healthy-salons/

Campaign for Safe Cosmetics. (n.d.) Available from: https://www.safecosmetics.org/

Dang, J., Rosemberg, M.-A., & Le, A. (2021). Perceived work exposures and expressed intervention needs among Michigan nail salon workers. International Archives of Occupational and Environmental Health, 94, 1–13. https://doi.org/10.1007/s00420-021-01719-6

Eggerth, D. E., & Flynn, M. A. (2010). When the Third World Comes to the First: Ethical Considerations When Working With Hispanic Immigrants. Ethics & Behavior, 20(3–4), 229–242. https://doi.org/10.1080/10508421003798968

Flynn, M. A., Check, P., Steege, A. L., Sivén, J. M., & Syron, L. N. (2021). Health Equity and a Paradigm Shift in Occupational Safety and Health. International Journal of Environmental Research and Public Health, 19(1), 349. https://doi.org/10.3390/ijerph19010349

Freeland, C., Huynh, T., Vu, N., Nguyen, T., & Cohen, C. (2021). Understanding Knowledge and Barriers Related to Hepatitis B for Vietnamese Nail Salon Workers in the City of Philadelphia and Some of Its Environs. Journal of Community Health, 46(3), 502–508. https://doi.org/10.1007/s10900-020-00878-w

Garcia, E., Sharma, S., Pierce, M., Bhatia, S., Argao, S. T., Hoang, K., & Quach, T. (2015a). Evaluating a County-Based Healthy Nail Salon Recognition Program. AMERICAN JOURNAL OF INDUSTRIAL MEDICINE, 58(2), 193–202. https://doi.org/10.1002/ajim.22379

Garcia, E., Sharma, S., Pierce, M., Bhatia, S., Argao, S. T., Hoang, K., & Quach, T. (2015b). Evaluating a County-Based Healthy Nail Salon Recognition Program. AMERICAN JOURNAL OF INDUSTRIAL MEDICINE, 58(2), 193–202. https://doi.org/10.1002/ajim.22379

Healthy Nail Salon Recognition Program. (n.d.). Department of Toxic Substances Control. Retrieved May 18, 2021, from https://dtsc.ca.gov/scp/healthy-nail-salon-recognition-program/

Healthy Nail Salons—Local Hazardous Waste Management Program in King County. (n.d.). Retrieved May 18, 2021, from https://www.hazwastehelp.org/health/nailsalonresources.aspx

Herrera, L., Waheed, S., Flowers, K., Fu, L., Nguyen, D., & Nguyen, C. (2020). A Survey of Nail Salon Workers and Owners in California During Covid-19. https://escholarship.org/uc/item/70b6t85q

Herrera, L., Waheed, S., Macia, M., Fu, L., Nguyen, T., Nguyen, D. (2021). The experience of nail salon owners and owners in California. California Healthy Nail Salon Collaborative and UCLA Labor Center. https://drive.google.com/file/d/1lNjjVO3RZu1PeWWZs2T6Wn9QHpziAUfL/view

Honan, J., Ingram, M., Quijada, C., Chaires, M., Fimbres, J., Ornelas, C., Sneed, S., Stauber, L., Spitz, R., Sandoval, F., Carvajal, S., Billheimer, D., Wolf, A. M., & Beamer, P. (2022). Understanding the Impacts of the COVID-19 Pandemic on Small Businesses and Workers Using Quantitative and Qualitative Methods. Annals of Work Exposures and Health, wxac048. https://doi.org/10.1093/annweh/wxac048

Huynh, T. B., Doan, N., Trinh, N., Verdecias, N., Stalford, S., & Caroll-Scott, A. (2019). Factors influencing health and safety practices among Vietnamese nail salon technicians and owners: A qualitative study. American Journal of Industrial Medicine, 62(3), 244–252. https://doi.org/10.1002/ajim.22947

Kononowech, J., Hagedorn, H., Hall, C., Helfrich, C. D., Lambert-Kerzner, A. C., Miller, S. C., Sales, A. E., & Damschroder, L. (2021). Mapping the organizational readiness to change assessment to the Consolidated Framework for Implementation Research. Implementation Science Communications, 2(1), 19. https://doi.org/10.1186/s43058-021-00121-0

LoSasso, G. L., Rapport, L. J., Axelrod, B. N., & Whitman, R. D. (2002). Neurocognitive sequelae of exposure to organic solvents and (meth)acrylates among nail-studio technicians. Neuropsychiatry, Neuropsychology, and Behavioral Neurology, 15(1), 44–55.

Ma, G. X., Wei, Z., Husni, R., Do, P., Zhou, K., Rhee, J., Tan, Y., Navder, K., & Yeh, M.-C. (2019). Characterizing Occupational Health Risks and Chemical Exposures Among Asian Nail Salon Workers on the East Coast of the United States. Journal of Community Health, 44(6), 1168–1179. https://doi.org/10.1007/s10900-019-00702-0

Morey, B. N. (2018). Mechanisms by Which Anti-Immigrant Stigma Exacerbates Racial/Ethnic Health Disparities. American Journal of Public Health, 108(4), 460–463. https://doi.org/10.2105/AJPH.2017.304266

Moutray C. (2009) Looking ahead: opportunities and challenges for entrepreneurship and small business owners. West New Engl Law Rev; 31: 763.

New York State. (n.d.). Nail salon safety: What you need to know. https://www.ny.gov/programs/nail-salon-safety-what-you-need-know

Nguyen, L. V., Diamond, M. L., Kalenge, S., Kirkham, T. L., Holness, D. L., & Arrandale, V. H. (2022). Occupational Exposure of Canadian Nail Salon Workers to Plasticizers Including Phthalates and Organophosphate Esters. Environmental Science & Technology. https://doi.org/10.1021/acs.est.1c04974

Quach, T., Nguyen, K.-D., Doan-Billings, P.-A., Okahara, L., Fan, C., & Reynolds, P. (2008). A preliminary survey of Vietnamese nail salon workers in Alameda County, California. Journal of Community Health, 33(5), 336–343. https://doi.org/10.1007/s10900-008-9107-7

Quiros-Alcala, L., Pollack, A. Z., Tchangalova, N., DeSantiago, M., & Kavi, L. K. A. (2019). Occupational Exposures Among Hair and Nail Salon Workers: A Scoping Review. CURRENT ENVIRONMENTAL HEALTH REPORTS, 6(4), 269–285. https://doi.org/10.1007/s40572-019-00247-3

Rodriguez-Lainz, A., McDonald, M., Fonseca-Ford, M., Penman-Aguilar, A., Waterman, S. H., Truman, B. I., Cetron, M. S., & Richards, C. L. (2018). Collection of Data on Race, Ethnicity, Language, and Nativity by US Public Health Surveillance and Monitoring Systems: Gaps and Opportunities. Public Health Reports, 133(1), 45–54. https://doi.org/10.1177/0033354917745503

Roelofs, C., Azaroff, L. S., Holcroft, C., Nguyen, H., & Doan, T. (2008). Results from a community-based occupational health survey of Vietnamese-American nail salon workers. J Immigr Minor Health, 10(4), 353–361. https://doi.org/10.1007/s10903-007-9084-4

Seller, S. L., Roelofs, C., Shoemaker, P. A., Nguyen, N. N., & Nguyen, T. D. (2019). Improving Boston Nail Salon Indoor Air Quality Through Local Public Health Regulation, 2007-2019. AMERICAN JOURNAL OF PUBLIC HEALTH, 109(12), 1711–1713. https://doi.org/10.2105/AJPH.2019.305329

Sharma, P., Gonzalez, L., Herrera, L., Waheed, S., Fu, L., Nguyen, D., Nguyen T., Gurung, P., Dash, P., Nguyen, N., & Tran, E. (2022). Working Under Covid-19: Experiences of Nail Salon. Adhikaar for Human Rights & Social Justice, California Healthy Nail Salon Collaborative, UCLA Labor Center and VietLead. https://drive.google.com/file/d/1lNjjVO3RZu1PeWWZs2T6Wn9QHpziAUfL/view?usp=sharing&usp=embed_facebook

Sharma, P., Waheed, S., Nguyen, V., Stepick L., Orellana, R., Katz, L., Kim, S., Lapira, K. (2018). Nail File: A Study of Nail Salon Workers and Industry in the United States. UCLA Labor Center and California Healthy Nail Salon Collaborative.

Siegel, M. R., Rocheleau, C. M., Broadwater, K., Santiago-Colón, A., Johnson, C. Y., Herdt, M. L., Chen, I.-C., & Lawson, C. C. (2021). Maternal occupation as a nail technician or hairdresser during pregnancy and birth defects, National Birth Defects Prevention Study, 1997–2011. Occupational and Environmental Medicine, oemed-2021-107561. https://doi.org/10.1136/oemed-2021-107561

The Consolidated Framework for Implementation Research – Technical Assistance for users of the CFIR framework. (n.d.). Retrieved January 11, 2022, from https://cfirguide.org/

U. S. Food and Drug Administration (2022). Nail Care Products. https://www.fda.gov/cosmetics/cosmetic-products/nail-care-products

Wallace, S. P., & Young, M. E. de T. (2018). Immigration Versus Immigrant: The Cycle of Anti-Immigrant Policies. American Journal of Public Health, 108(4), 436–437. https://doi.org/10.2105/AJPH.2018.304328

